# Deciphering microbiota of acute upper respiratory infections: a comparative analysis of PCR and mNGS methods for lower respiratory trafficking potential

**DOI:** 10.1101/2023.01.03.22284078

**Authors:** Sadia Almas, Rob E. Carpenter, Anuradha Singh, Chase Rowan, Vaibhav K. Tamrakar, Rahul Sharma

## Abstract

Although it is clinically important for acute respiratory tract (co)infections to have rapid and accurate diagnosis, it is critical that respiratory medicine understand the advantages of current laboratory methods. In this study we tested nasopharyngeal samples (n=29) with a commercially available PCR assay and compared the results with hybridization capture-based mNGS workflow. Detection criteria for positive PCR samples was Ct<35 and for mNGS samples >40% target coverage, median depth of 1X and RPKM>10. A high degree of concordance (98.33% PPA and 100% NPA) was recorded. However, mNGS yielded positive 29 additional microorganisms (23 bacteria, 4 viruses, and 2 fungi) beyond PCR. We then characterize the microorganisms of each method into three phenotypic categories using the IDbyDNA Explify® Platform for consideration of infectivity and trafficking potential to the lower respiratory region. The findings are significant for providing a comprehensive yet clinically relevant microbiology profile of acute upper respiratory infection, especially important in immunocompetent respiratory cases or where traditional syndromic approaches fail to identify pathogenicity. Accordingly, this technology can be used to supplement current syndromic-based tests and data can quickly and effectively be phenotypically characterized for trafficking potential, clinical (co)infection, and comorbid consideration—with promise to reduce morbidity and mortality.

**Highlights:** *What are the main findings?:*

- Although there was a high concordance between methodologies, a hybridization capture-based mNGS workflow was able to detect 29 additional upper respiratory microorganisms versus PCR.
- Identified microorganisms were rapidly characterized into three phenotypic groups for infectivity and trafficking potential.

*What is the implication of the main finding?:*

- A hybridization capture-based mNGS workflow can provide a comprehensive yet clinically relevant microbiology profile of acute upper respiratory infection
- Deciphering upper respiratory microbiota with phenotypic grouping has potential to provide respiratory medicine a tool to better manage immunocompetent and complex respiratory cases.

## 1. Introduction

Preceding or concurrent upper respiratory (co)infections can have harmful trafficking effects on lower respiratory disease. And lower respiratory (co)infections are a communal source of worldwide morbidity and mortality [1]. The clinical picture of human upper and lower respiratory infections can be complex and heterogenous since etiological agents (i.e., bacteria, fungi, viruses, and parasites) can be present alone or in combination. For example, the consequences of viral-bacterial (co)infections have been increasingly recognized for affecting the manifestation and prognosis of community-acquired pneumonia and can profoundly impact concomitant development of respiratory disease, frequently resulting in the need for intensive care [2-6]. Especially at risk are children under the age of 1, pregnant women, the elderly, and immunocompromised hosts. Immunocompetent individuals with comorbid illness are also at increased risk of severe respiratory infection often requiring intensive care [7]. The recent COVID-19 pandemic further highlighted that viral performance often has devastating effects on human health when coupled with fungal and bacterial (co)infections [8]. No doubt, the burden of respiratory (co)infection is a major threat to global health and the need for timely and accurate diagnosis is universal [9,10].

It is clinically important for acute respiratory tract (co)infections to have rapid and accurate diagnosis to reduce risk of protracted (co)infections and advance application of pathogen specific medication—taking into account the worsening universal problem of antibiotic resistant microbes [11,12]. For example, multiplex polymerase chain reaction (PCR) assays have advanced diagnosis for numerous respiratory pathogens and antimicrobial resistance (AMR) markers in a single panel, reducing diagnosis time and by-passing other serial tests like serology and culture to identify respiratory (co)infections [13]. However, it is critical to understand these syndromic approaches have limitations rooted in techniques based upon *a priori* assumptions and a validated scope of target specific agents; meaning these methods are biased to a set of predetermined microbes with limited capacity to discover or differentiate clinically relevant strains or genotypes [9,14]. As a result, these traditional laboratory approaches may not identify (co)infectivity, trafficking potential, fastidious microorganisms, rare and atypical pathogens, or agents inviable on culture post antimicrobial therapy.

On the other end of the spectrum, the target agnostic approach of metagenomic next-generation sequencing (mNGS) technology has potential to accurately identify respiratory (co)infections without *a priori* knowledge aimed at broadening pathogen discovery, shortening detection time for certain microorganisms, and with detection strength less affected by past antibiotic exposure. This hypothesis-free approach has emerged as a promising laboratory method; yielding higher pathogen identification, uncovering progressive (co)infections, and for directly influencing patient care—including suitable antibiotic coverage and reduced mechanical ventilation [15-17]. And one of the greatest attributes of mNGS may be its ability to capture a patient’s microbiome to detect comorbid infections that may complicate treatment and recovery [18] But mNGS, too, has limitations that are critical to understand for respiratory medicine. Because of the target agnostic approach, mNGS data has potential to query all microbiome in the sample; the significance in acquiring thousands to millions of short DNA sequences from a single respiratory sample can be a taxonomic burden and relies primarily on being able to resolve clinically relevant data. In doing so, several challenges exist in experimental design and computational analysis between pathogenicity and the true microbiome in samples such as respiratory fluid [19]—with interpretation of the data for case relevance in respiratory medicine another matter altogether.

For these reasons, targeted (precision) metagenomics—a hybridization capture-based mNGS approach—is becoming more commonly considered for targeted sequencing in clinical settings [20-23]. Hybridization capture is especially helpful because it uses biotinylated oligonucleotide probes to focus on specific genomic regions of interest. Accordingly, its multiplexing capacity is improved by enabling ‘molecular barcodes’ that can be ligated, combined, and pooled with several samples at equal mass, reducing workflow effort and cost. Moreover, a targeted hybridization capture-based mNGS approach can maximize on-target reads, improve mutation exactitude, and provide superior performance with complex sequences, making it an especially appealing option for analyzing respiratory infections. And although researchers have demonstrated that such hybridized workflows can provide critical pathogen-specific sequences for lower respiratory (co)infections [24], preceding or concurrent consideration of upper respiratory (co)infections with potential to orchestrate specific leukocyte trafficking molecules to inform a course of lower respiratory pathology remains an open challenge in the fields of laboratory and respiratory medicine [25].

The purpose of this study is twofold. First, this research helps to extend our understanding of the potential clinical utility of a hybridization capture-based mNGS workflow for respiratory medicine by deploying a Respiratory Pathogen ID/AMR enrichment panel (RPIP) (187 bacteria, 42 viruses, 53 fungi, and 1,218 AMR; See https://www.illumina.com/products/by-type/sequencing-kits/library-prep-kits/respiratory-pathogen-id-panel.html for additional information) to test nasopharyngeal samples (*n* = 29) of individuals suspected of acute upper respiratory infection and comparatively analyze the results of identical samples using the Fast Track Diagnostic (FTD®) Respiratory Pathogens 33 (RUO) PCR panel (21 viruses, 12 bacteria, 1 fungus, and 2 AMR). Second, we characterize the discovered microorganisms of each method into phenotypic categories using the IDbyDNA Explify® Platform for consideration of trafficking concerns to the lower respiratory region for the attention of respiratory medicine.

## 2. Materials and Methods

### 2.1. Nucleic acid isolation

Nucleic acid was extracted from nasopharyngeal samples (*n* = 29) collected from individuals recommended for respiratory pathogen PCR testing at Advanta Genetics (Texas, USA). Two different methods were used for nucleic acid extraction for PCR and mNGS analysis; total nucleic acid was extracted for PCR analysis, and DNA and RNA were extracted separately for mNGS analysis. Briefly, total nucleic acid isolation was performed as part of routine diagnostic testing using the MagNA Pure 96 (MP96) DNA and Viral NA Small Volume Kit (Cat # 06543588001). Briefly, samples were lysed with 340 uL of lysis buffer and 10 uL of proteinase K (Invitrogen Cat # 4333793) at 55°C for 10 minutes, followed by extraction via the MP96 instrument. Extracted nucleic acids were stored at -80°C until used for PCR testing.

DNA and RNA from each sample were extracted separately using the Zymo research reagents as per protocol provided in the Explify Respiratory Pathogen ID/AMR Panel User Guide (Illumina Inc/IDbyDNA, 2022, P. 10). Each sample was spiked with T7 bacteriophage DNA (Microbiologics, St. Cloud, MN), delivering a final concentration of 1.2 × 10^7^ plaque forming units (PFU/mL) of sample. Copies of T7 were used for computing the absolute concentration of the target copies detected in the samples. Briefly, 400ul of the sample was homogenized and lysed by vertexing in ZR Bashing Beads (Cat# S6012-50). Homogenate supernatant was mixed with DNA/RNA Lysis Buffer (Cat # D7001-1), and DNA was first extracted using the Spin-Away Filter (Cat #. C1006). Flow-through from the DNA extraction was used for RNA extraction using the Zymo-Spin IIC (Cat #. C1011). RNA was treated with the Zymo DNases I enzyme. (Cat # E1009-A).

### 2.2. PCR testing using Fast Track Diagnostic® assay

Real-time PCR testing was performed using TaqMan chemistry based Fast Track Diagnostic® (FTD) Respiratory Pathogens 33 (RUO) kit. Ten microliter multiplex reactions targeting 13 bacteria, 19 viruses, and one fungal pathogen were performed in 384 well plate format on a Light Cycler® 480 System (Roche) instrument (Supplementary Table-1). One step reverse transcription PCR was performed in 11 multiplex PCR reactions, and each reaction was targeted to detect three pathogens. The multiplex real time RT-PCR thermal cycling profile for the FTD kit includes cDNA synthesis at 50 °C for 15 min, and initial denaturation at 95 °C for 10 min followed by 40 cycles of PCR amplification at 95 °C for 8 sec and 60 °C for 34 sec. PCR results were considered positive for the targets if threshold cycle (Ct) values were ≤35 paired with sigmoidal amplification curves.

### 2.3. Library preparation and enrichment

Sequencing libraries were prepared for the mNGS using Illumina®/IDbyDNA Respiratory Pathogen ID/AMR Panel (RPIP) protocol and reagents (Illumina® Inc, USA). Briefly, cDNA was prepared from the RNA and combined with the DNA in equal volumes. Libraries were constructed by DNA tegmentation and adapter ligation using the Illumina® RNA Prep with enrichment kit. Libraries were enriched for the microbial content by hybridization with the RPIP probes for 2 hrs. Captured libraries were amplified for 14 cycles and cleaned using AmPure XP beads (Beckman Coulter, Inc. USA). Two NATtrolTM Respiratory Panel 2.1 (RP2.1) Controls (Cat# NATRPC2.1-BIO) (ZeptoMetrix Inc. USA) and a blank viral transport medium (VTM) (Criterion Chemistries AL, USA) were included as positive and negative controls respectively with each batch of library preparation and sequencing. Libraries were quantified using a Qubit Flex Fluorometer (Thermo Fisher Scientific Inc USA) and fragment sizes of representative libraries were analyzed in Agilent 5200 Fragment Analyzer. The enriched libraries were then pooled to an equimolar concentration and normalized to 1nM concentration. The final library pool was denatured and neutralized with 0.1 N NaOH and 200 mM Tris-HCL (pH-8). The denatured libraries were further diluted to a loading concentration of 2 particulate matter. Dual indexed paired-end sequencing with 75 bp read length was performed using the high-output flow cell (150 cycles) on the Illumina MiniSeq® instrument. Although the goal depth for this workflow was 1.0 million reads, samples with 0.5 million reads were included in the downstream analysis.

### 2.4. Bioinformatic analysis using the Explify® Platform

Sequencing data were analyzed using the automated IDbyDNA Explify® Platform data analysis solution (v1.0.1). This software detects 282 pathogens, covering more than 95% of common and rare pathogens of respiratory infections. This software also detects 1218 AMR markers to predict the resistance of 12 common bacterial pathogens to 16 commonly used antibiotics. However, AMR analysis was not included in this comparative study.

Following analysis, a report was generated that included a detailed text-based (JSON format) and .pdf document that contained the quantitative identification of viruses, bacteria, and fungus in each sample, including the AMR markers. Each identified microorganism was assigned to phenotypic groups (1, 2 & 3) based on its potential pathogenic status. Group 1 microorganisms are frequently considered part of the normal flora but may be associated with disease in certain settings. Microorganisms in group 2 are frequently associated with the disease, and group 3 microorganisms are generally considered to be associated with the disease. Accurate detection of the known microorganisms in positive controls was used for defining the acceptance criteria for target detection in clinical samples.

## 3. Results

Identical nasopharyngeal samples (n=29) collected from individuals with suspected acute upper respiratory infections were tested using a syndromic PCR panel (Supplementary Table 1) and a hybridization capture-based mNGS workflow (Supplementary Table 2). All data were analyzed qualitatively. Serial dilutions of Zeprometrix respiratory controls (1 and 2) were tested with each batch of the clinical samples and only controls with >0.5million total reads were resulted in accurate detection of all included targets. Thus, samples not resulted in minimum of 0.5million reads were excluded from the further analysis. Further, the minimum coverage and reads per kilobase per million reads mapped (RPKM) required for >90% accurate detection of the targets in controls was used as acceptance criteria for the microorganism detection in clinical sample results. Detection criteria for positive microorganisms were Ct<35 in the PCR assay, and >40% target coverage, median depth of 1X and RPKM>10 in the mNGS assay. Results from the two methodologies were comparatively analyzed and phenotypically characterized into groups according to their infectivity potential.

### 3.1. Bioinformatic analysis using the Explify® Platform

Analyte-specific PCR analysis detected 28/29 samples positive for one or more microorganism(s). One sample was negative for all tested microorganisms. Overall, 15 etiological agents (4 bacteria and 11 viruses) were identified among the 28 positive samples tested by PCR (Table 1). Analysis of the 28 positive samples revealed 9 samples exclusively positive for bacteria and 9 samples exclusively positive for viruses, whereas 10 samples were concomitantly positive for both bacteria and viruses. The most common microorganisms detected by PCR were *Moraxella catarrhalis, Hemophilus influenza*, and *Streptococcus pneumoniae*.

**Table 1.** Fast Track

| FTD® PCR Panel | Microorganism Classification | PCR Positive |
| --- | --- | --- |
| *Moraxella catarrhalis* | Bacteria | 13(45%) |
| *Haemophilus influenzae* | Bacteria | 9(31%) |
| *Streptococcus pneumoniae* | Bacteria | 9(31%) |
| *Staphylococcus aureus* | Bacteria | 5(17%) |
| Human rhinovirus | Virus | 4(14%) |
| Influenza A virus (H3N2) | Virus | 4(14%) |
| Human coronavirus OC43 | Virus | 3(10%) |
| Human metapneumoviruses | Virus | 3(10%) |
| Human respiratory syncytial viruses | Virus | 3(10%) |
| Human parainfluenza 1 virus | Virus | 2(7%) |
| Enterovirus | Virus | 1(3%) |
| Human parainfluenza 2 virus | Virus | 1(3%) |
| Human parainfluenza 3 virus | Virus | 1(3%) |
| Human parainfluenza 4 virus | Virus | 1(3%) |

Diagnostic® (FTD) assay results (n = 29)

The phenotypic grouping of positive microorganisms was then classified according to their infectivity potential: 0/28 (0%) microorganisms were phenotypically classified (phenotypic group 1) as part of the normal flora, colonizers, or contaminants; 3/28 (11%) microorganisms were phenotypically classified (phenotypic group 2) as frequently associated with respiratory disease; and 12/28 (43%) microorganisms were phenotypically classified (phenotypic group 3) as pathogenic for respiratory disease.

### 3.2. Bioinformatic analysis using hybridization capture-based mNGS workflow

The hybridization capture-based mNGS workflow used in this study probed an additional 249 microorganisms (23 viruses, 174 bacteria, and 52 fungi) that were not included in the PCR panel. However, only samples yielding >0.5 million reads were included in the downstream mNGS analysis; microorganisms with coverage ≥40.00%, median depth ≥1X and RPKM ≥10.00 were considered positive. The hybridization capture-based mNGS workflow identified 44 microorganisms (27 bacteria, 2 fungi, and 15 viruses) in 29 samples (Table 2; Figure 1).

**Table 2.** Illumina® RPIP mNGS panel positive microorganisms

| Illumina® RPIP mNGS Panel | Microorganism Classification | mNGS Positive |
| --- | --- | --- |
| *Moraxella catarrhalis* | Bacteria | 12(41%) |
| *Dolosigranulum pigrum* | Bacteria | 12(41%) |
| *Haemophilus influenzae* | Bacteria | 9(31%) |
| *Streptococcus pneumoniae* | Bacteria | 9(31%) |
| *Stenotrophomonas maltophilia* | Bacteria | 7(24%) |
| *Pseudomonas aeruginosa* | Bacteria | 6(21%) |
| *Staphylococcus aureus* | Bacteria | 5(17%) |
| *Corynebacterium pseudodiphtheriticum* | Bacteria | 4(14%) |
| Human rhinovirus A | Virus | 4(14%) |
| Influenza A virus (H3N2) | Virus | 4(14%) |
| *Corynebacterium propinquum* | Bacteria | 3(10%) |
| Human coronavirus OC43 | Virus | 3(10%) |
| Human metapneumovirus | Virus | 3(10%) |
| *Ochrobactrum anthropi* | Bacteria | 3(10%) |
| *Prevotella melaninogenica* | Bacteria | 3(10%) |
| Respiratory syncytial virus B | Virus | 3(10%) |
| *Rothia mucilaginosa* | Bacteria | 3(10%) |
| *Streptococcus mitis* | Bacteria | 3(10%) |
| *Actinomyces graevenitzi* | Bacteria | 2(7%) |
| *Alternaria alternata* | Fungus | 2(7%) |
| *Campylobacter concisus* | Bacteria | 2(7%) |
| *Capnocytophaga leadbetteri* | Bacteria | 2(7%) |
| Cytomegalovirus (CMV) | Virus | 2(7%) |
| *Gemella haemolysans* | Bacteria | 2(7%) |
| Human parainfluenza virus 1 | Virus | 2(7%) |
| SARS-CoV-2 (2019-nCoV) | Virus | 2(7%) |
| *Veillonella parvula* | Bacteria | 2(7%) |
| *Achromobacter xylosoxidans* | Fungus | 1(3%) |
| *Actinomyces naeslundii* | Fungus | 1(3%) |
| Coxsackievirus A | Virus | 1(3%) |
| Enterovirus D68 | Virus | 1(3%) |
| *Fusarium proliferatum* | Fungus | 1(3%) |
| *Fusobacterium necrophorum* | Fungus | 1(3%) |
| *Haemophilus haemolyticus* | Bacteria | 1(3%) |
| *Haemophilus parainfluenzae* | Bacteria | 1(3%) |
| Human parainfluenza 4 virus | Virus | 1(3%) |
| Human parainfluenza virus 2 | Virus | 1(3%) |
| Human parainfluenza virus 3 | Virus | 1(3%) |
| Human rhinovirus C | Virus | 1(3%) |
| Influenza A virus (H1N1) | Virus | 1(3%) |
| *Neisseria flavescens* | Bacteria | 1(3%) |
| *Neisseria lactamica* | Bacteria | 1(3%) |
| *Pseudomonas stutzeri* | Bacteria | 1(3%) |
| *Streptococcus intermedius* | Bacteria | 1(3%) |

**Figure 1.**
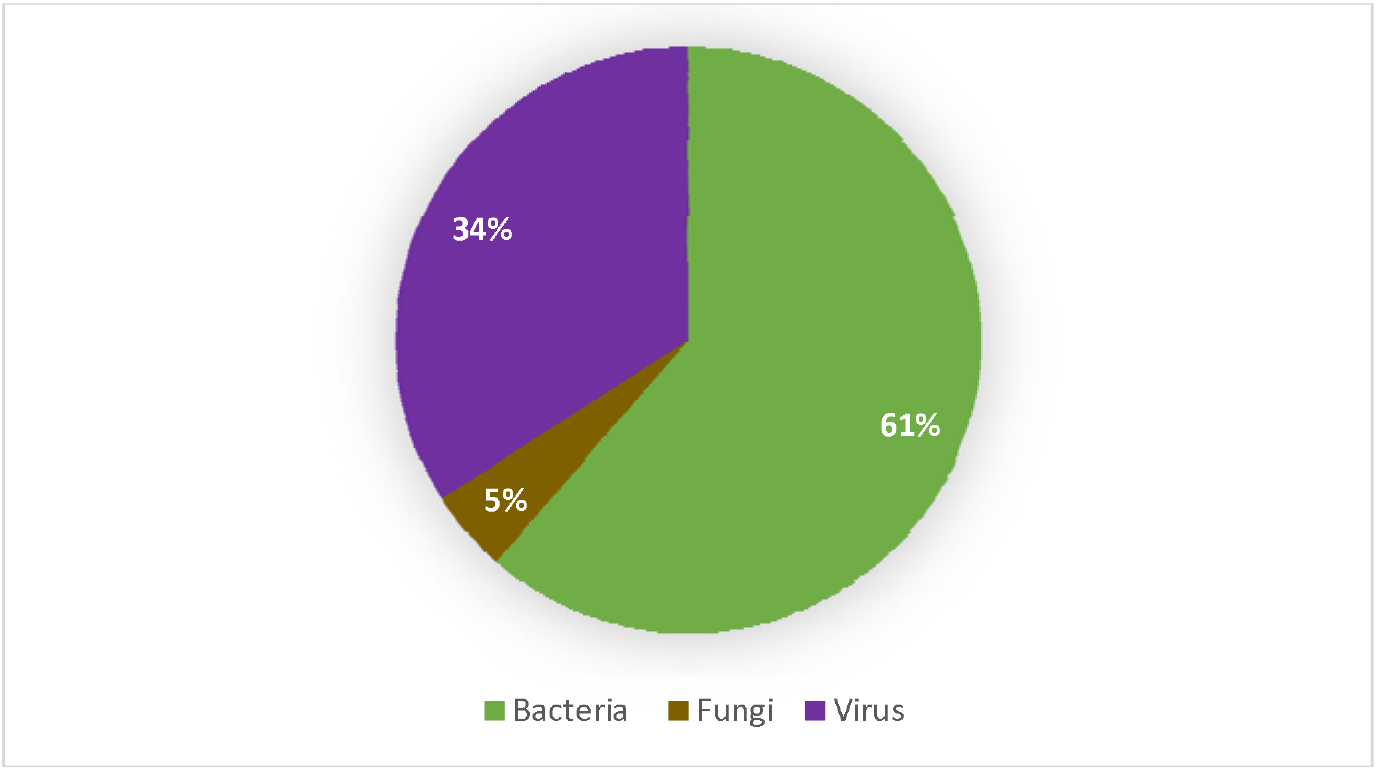
Classification of microorganisms detected by mNGS (coverage ≥40.00%, median depth ≥1X and RPKM ≥10.00)

The results demonstrated only 3 samples were positive for a single organism (S. aureus, HRV-C, A/H1N1), 5 samples were exclusively co-infected with bacterial species, and 2 samples were positive for multi-viral infection. Bacterial and viral coexistence was the most common presentation identified (n = 16), followed by bacterial and fungal coexistence in 2 samples. Only one sample was found concurrently colonized with bacteria, viruses, and fungus. The phenotypic grouping of positive microorganisms were then classified according to their infectivity potential: 14/44 (31%) microorganisms were phenotypically classified (phenotypic group 1) as part of the normal flora, colonizers, or contaminants; 15/44 (34%) microorganisms were phenotypically classified (phenotypic group 2) as frequently associated with respiratory disease; and 15/44 (34%) microorganisms were phenotypically classified (phenotypic group 3) as pathogenic for respiratory disease.

### 3.3. Comparative analysis between PCR and mNGS assays

The results of both assays were analysed, compared, and phynotypically classified (Table 3). A high degree of concordance (98.33% PPA and 100% NPA) was recorded between the PCR and mNGS results for the targets (n = 33) shared by both panels. Only one sample showed discordance when M. catarrhalis was detected by PCR and not mNGS. While 4 bacteria and 11 viruses were concurrently detected by the PCR and mNGS panels, mNGS yielded positive for 29 additional microorganisms (23 bacteria, 4 viruses, and 2 fungi) beyond the PCR panel. Granted, the mNGS panel identified the colonization of a wide range of upper respiratory tract flora with (14/44; 13 bacterial, 1 fungi) that were unlikely pathogenic and classified to phenotypic group 1. Among the 15 microorganisms detected and assigned to phenotypic group 2, mNGS analysis exclusively detected 12/15—these microorganisms are frequently associated with lower respiratory disease and detection of these microorganisms is potentially clinically significant. Coxsackievirus A, SARS-CoV-2 (2019-nCoV) and HRV-C were exclusively identified by mNGS and classified to phenotypic group 3 as pathogenic with Explify® analysis. The two samples found positive for SARS-CoV-2 and were separately tested with SARS-CoV-2 specific PCR assays; both were confirmed positive. The remaining 12 organisms assigned to phenotypic group 3 (generally considered disease-associated) were in parallel with the PCR panel and were concurrently detected by both assays (Figure 2).

**Table 3.**
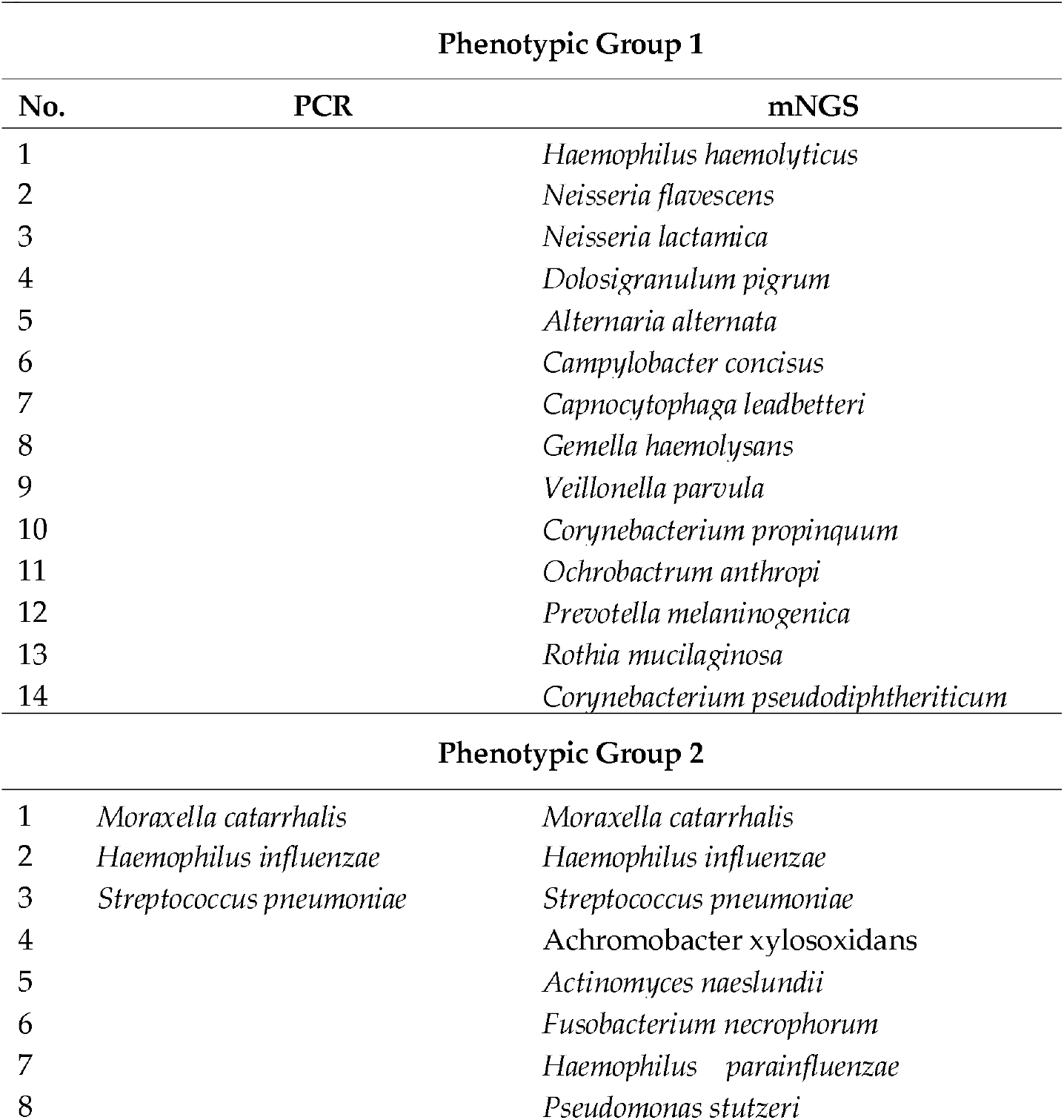
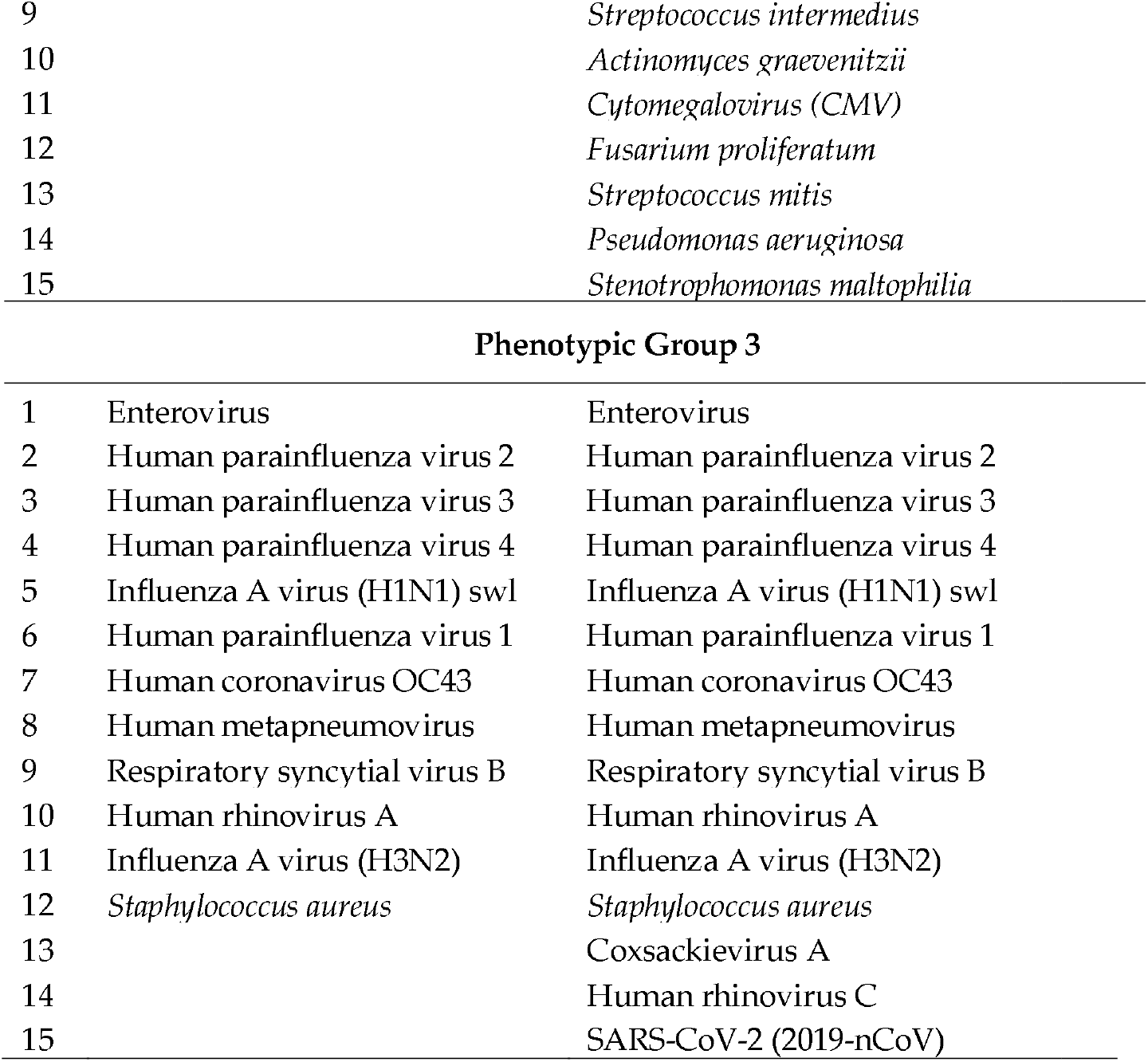
PCR vs mNGS phenotypic grouping of upper respiratory tract microorganisms

| Phenotypic Group 1 |  |  |
| --- | --- | --- |
| No. | PCR | mNGS |
| 1 |  | *Haemophilus haemolyticus* |
| 2 |  | *Neisseria flavescens* |
| 3 |  | *Neisseria lactamica* |
| 4 |  | *Dolosigranulum pigrum* |
| 5 |  | *Alternaria alternata* |
| 6 |  | *Campylobacter concisus* |
| 7 |  | *Capnocytophaga leadbetteri* |
| 8 |  | *Gemella haemolysans* |
| 9 |  | *Veillonella parvula* |
| 10 |  | *Corynebacterium propinquum* |
| 11 |  | *Ochrobactrum anthropi* |
| 12 |  | *Prevotella melaninogenica* |
| 13 |  | *Rothia mucilaginosa* |
| 14 |  | *Corynebacterium pseudodiphtheriticum* |
| Phenotypic Group 2 |  |  |
| 1 | *Moraxella catarrhalis* | *Moraxella catarrhalis* |
| 2 | *Haemophilus influenzae* | *Haemophilus influenzae* |
| 3 | *Streptococcus pneumoniae* | *Streptococcus pneumoniae* |
| 4 |  | *Achromobacter xylosoxidans* |
| 5 |  | *Actinomyces naeslundii* |
| 6 |  | *Fusobacterium necrophorum* |
| 7 |  | *Haemophilus parainfluenzae* |
| 8 |  | *Pseudomonas stutzeri* |

|  |  |
| --- | --- |
| 9 | *Streptococcus intermedius* |
| 10 | *Actinomyces graevenitzii* |
| 11 | *Cytomegalovirus (CMV)* |
| 12 | *Fusarium proliferatum* |
| 13 | *Streptococcus mitis* |
| 14 | *Pseudomonas aeruginosa* |
| 15 | *Stenotrophomonas maltophilia* |
| Phenotypic Group 3 |  |  |
| 1 | Enterovirus | Enterovirus |
| 2 | Human parainfluenza virus 2 | Human parainfluenza virus 2 |
| 3 | Human parainfluenza virus 3 | Human parainfluenza virus 3 |
| 4 | Human parainfluenza virus 4 | Human parainfluenza virus 4 |
| 5 | Influenza A virus (H1N1) swl | Influenza A virus (H1N1) swl |
| 6 | Human parainfluenza virus 1 | Human parainfluenza virus 1 |
| 7 | Human coronavirus OC43 | Human coronavirus OC43 |
| 8 | Human metapneumovirus | Human metapneumovirus |
| 9 | Respiratory syncytial virus B | Respiratory syncytial virus B |
| 10 | Human rhinovirus A | Human rhinovirus A |
| 11 | Influenza A virus (H3N2) | Influenza A virus (H3N2) |
| 12 | *Staphylococcus aureus* | *Staphylococcus aureus* |
| 13 |  | Coxsackievirus A |
| 14 |  | Human rhinovirus C |
| 15 |  | SARS-CoV-2 (2019-nCoV) |

**Figure 2.**
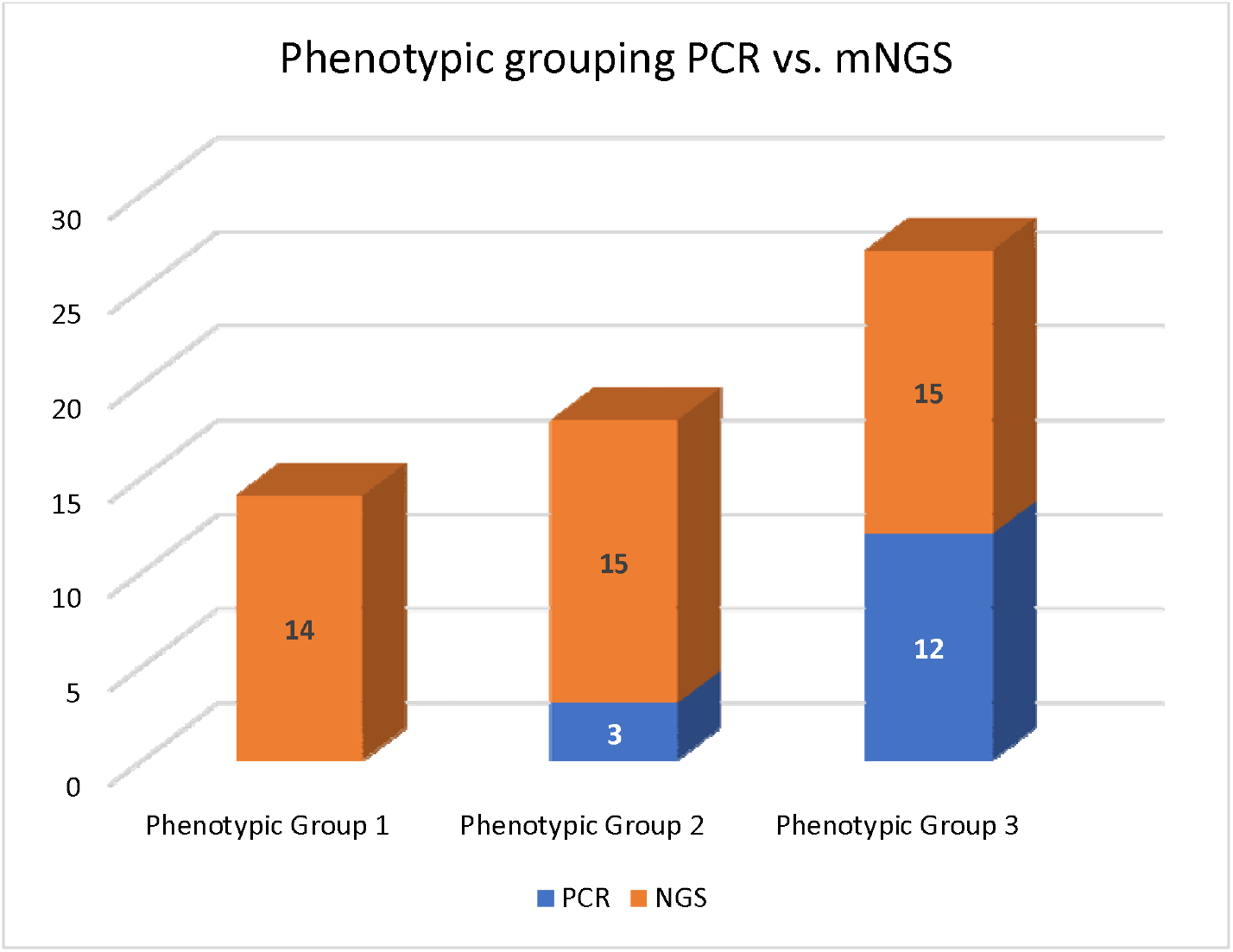
Comparative analysis of phenotypic grouping (PCR vs mNGS; n = 29)

## 4. Discussion

Respiratory (co)infections remain a leading cause of global morbidity and mortality despite advances in diagnosis and treatment. Rapid identification and functional characterization of microorganisms is critical for the clinical management of respiratory disease. No doubt, improved diagnostics of complex upper respiratory (co)infections is key to understanding the trafficking potential to the lower respiratory tract and their pathogenic role in lower airway disease [25]. However, there are noted limitations with traditional syndromic approaches to pathogen detection (i.e., serology, microbiology, PCR) that can result in lag-time to diagnosis, misdiagnosis, or unsuitable treatment, which increases risk for trafficking and prolonged sickness [26]. Important because etiology of respiratory tract (co)infections goes undiagnosed in ∼20 to 60% of patients with community-acquired pneumonia when using traditional syndromic laboratory testing approaches [14]. And more robust mNGS hypothesis-free techniques, although yielding a broad microbiome, pose a taxonomic-to-clinically relevant challenge. From a laboratory point of view these sequencing methods often require workflows that are technically challenging and tend to be siloed [27] because of the high capital investments needed for equipment, technology, and expertise—resulting in restricted capability for decentralized testing or application in resource-limited laboratories [14,28]. Yet from the clinical point of view, recognition of potential pathogenicity is crucial to estimate etiological relevance, to optimize treatment, and understand outbreak conditions. Similarly, the recognition of broader pathogenic microorganisms can be pivotable to draw on for decentralized epidemiological layouts [29]. Assuredly, underdiagnosed upper respiratory (co)infections have potential for trafficking to the lower respiratory region and trigger adverse harm, especially in immunosuppressed hosts [30-33].

More recent advances have introduced a hybridization capture-based mNGS approach with some initial success as a viable alternative for deciphering microbiota in clinical samples suspected of lower respiratory (co)infections [24]. However, exploring acute upper respiratory (co)infections with this approach to account for potential trafficking to the lower respiratory tract is less studied. Accordingly, we sought to test nasopharyngeal samples of patients suspected of upper respiratory (co)infection and (a) compare the findings of a commercially available respiratory PCR panel with a hybridization capture-based mNGS respiratory panel, and (b) phenotypically classify the findings for potential trafficking pathogenicity for the fields of laboratory and respiratory medicine to consider.

Second, the findings were characterized and phenotypically grouped using the automated IDbyDNA Explify® Platform data analysis solution (Tables 7-9). Of the normal flora, colonizers, or contaminants noted in phenotypic group 1, most are innocuous, commonly existing in commensal relationships with their hosts, having rare or low trafficking potential for active disease. Indeed, however, there are cases in literature to be considered. For example, bacteria such as *H. haemolyticus* has potential for a rare invasive disease which can be overlooked and misidentified as *H. influenzae* [35]; *N. flavescens* has been identified to have a pathogenic role in immunocompromised and diabetic patients, and in rare cases has been linked to necrotizing pneumonia [36]; *D. pigrum* has been shown to trigger cases of nosocomial pneumonia [37]; *C. leadbetteri* is occasionally responsible for acute exacerbation of chronic obstructing pulmonary disease (COPD) [38]; *G. haemolysans* has been discovered to play a direct role in pulmonary exacerbations in patients with cystic fibrosis [39]; although considered lower virulence, *Ochrobactrum* spp. are beginning to emerge in the literature as a major opportunistic respiratory pathogen [40]; *P. melaninogenica* has been found association with ventilator-associated pneumonia [41]; *R. mucilaginosa* has been linked to bronchiectasis [42]; and *C. pseudodiphtheriticum* has been associated with tracheobronchitis, pneumonia and lung abscesses [43], while others have shown trafficking potential to that includes various systemic infections, including meningitis and osteomyelitis [44]. Even more rare, but in fact with clinical manifestation, is the fungus *A. alternata*, which has been shown to have opportunistic infectivity in the lower respiratory tract in patients with acquired immunodeficiency syndrome [45].

In phenotypic group 2 we detected microorganisms more likely to persuade lower respiratory tract infectivity—frequently associated with respiratory disease. First, we consider three bacteria detected in parallel by both assays, *M. catarrhalis* (*n*=12/29), *H. influenzae* (*n*=9/29), and *S. pneumoniae* (*n*=9/29). Each of these bacteria has moderate to significant lower respiratory trafficking capacity. *M. catarrhalis*, although commonly noted as a cause of otitis media in children, has potential for COPD exacerbations and pathogenesis for bronchopulmonary infection post pulmonary aspiration [46,47]. Of course, *H. influenzae* and *S. pneumoniae* are common trafficking infectors originating in the upper respiratory tract. But even with vaccines, ∼1000 people die annually of *H. influenzae* in the United States, whereas *S. pneumoniae* is the leading cause of global pneumonia mortality [48]—confirming the importance of differentiating these pathogens on any targeted laboratory testing application. Regarding the remaining 12 microorganisms characterized as phenotypic group 2 (10 bacteria, 1 virus, 1 fungus; Table 7), the literature considers each associated with morbidity and mortality in immunocompromised and enervated individuals. Of more specific concern for trafficking potential is *H. parainfluenzae*, a commensal upper respiratory microorganism that with the acquisition of transmitted virulence can trigger a severe pathogen process in the lower respiratory tract [49]; *A. graevenitzii* as a cause of lung abscesses mimicking acute pulmonary coccidioidomycosis [50]; *A. xylosoxidans* for its predilection to worsen certain cases of cystic fibrosis [51]; and *S. mitis* and *S. maltophilia* due to their recognition as increasingly significant nosocomial pathogens that frequently exhibit multidrug resistance [52,53]. In addition to the identified bacteria, CMV spp. was found in 2/29 cases and has potential for severe symptomatic pneumonia in immunocompetent hosts [54] and F. proliferatum was observed in 1/29 cases, critical for its infectivity potential in lung transplant patients [55].

Phenotypic group 3 microorganisms are considered for their more significant pathogenicity aimed at respiratory disease. Importantly, 12/15 microorganisms (Table 9) were identified in parallel between PCR and mNGS—again confirming the significance of clinically relevant PCR targets in the laboratory. However, the hybridization capture-based mNGS workflow discovered three pathogens not probed by PCR, each with significant infectivity potential for trafficking to the lower respiratory tract. First, SARS-CoV-2, the responsible agent for the COVID-19 pandemic with global deaths of 6.5 million and climbing [56]. The importance of including a probed target for SARS-CoV-2 in any respiratory case or laboratory test cannot be understated, and we leave this discussion to scholars who have written much on this topic [57-59]. We do, however, consider in more depth the additional two microorganisms exclusively identified by mNGS that were characterized in this group: CAV and HRV-C—both targeting bronchial and alveolar epithelial cells and explicit for excessive uncontrolled lung inflammation if not cleared. When considering these pathogens, it is important for clinicians to take into account most PCR panels, including the FTD® panel used in this study, use primers that do not distinguish between enterovirus and rhinovirus, and report positive amplification without any enterovirus/rhinovirus genus differentiation. While targeted PCR has tremendous advantages that include cost, scale, speed, sensitivity and specificity, the results are restricted in divergence, diversity, genotype, and functional potential, ensuing in restricted clinical significance in many cases of respiratory infection [60,61].

Important because the genus *Enterovirus* has been divided into a total of 12 species; enteroviruses A-J (which include the coxsackievirus, poliovirus, and echovirus subspecies) and Human rhinoviruses A-C. Coxsackie viruses— reactivating pathogens in immunosuppressed patients—are divided into two major serotype groups: group A and group B. And although type A coxsackieviruses cause Herpangina (a common childhood illness), acute hemorrhagic conjunctivitis, and hand-foot-and-mouth disease, there are case reports—from toddler to adult—of fatal pulmonary illnesses as a result of CAV [62,63]. Of the three Human rhinoviruses A-C, HRV-C, identified in 2006, is more frequently associated with its severity of clinical manifestations for lower respiratory tract disease and severe illness [64,65]. For this reason, hybridization capture-based mNGS workflow may have clinical significance for its ability to differentiate these viruses at the species level—especially valuable in pediatric and immunocompromised cases {66,67].

### 4.1 Implications for respiratory medicine

Because lower respiratory (co)infections are a communal source of worldwide morbidity and mortality [1], there is a need to identify preceding or concurrent upper respiratory (co)infections that may trigger harmful trafficking effects on lower respiratory disease. And because the clinical picture of lower respiratory (co)infections can be complex and heterogenous with multiple etiological agents (i.e., bacteria, fungi, viruses, and parasites), rapid and accurate diagnosis can reduce risk of protracted (co)infections and advanced application of pathogen specific medication. However, when considering current syndromic approaches to diagnosis, respiratory medicine must consider limitations in laboratory testing rooted in techniques based upon *a priori* assumptions. In such cases, where accurate identification of diagnosis of fastidious microorganisms, rare and atypical pathogens, or inviable agents post antimicrobial therapy, a hybridization-capture-based mNGS approach may offer a viable alternative.

This is because clinical samples often comprise nucleic acids derived from the host (human), whereas the nucleic acids of interest for a hybridization-capture-based mNGS approach are microbial (or nonhuman). Thus, hybridization-capture-based enrichment can secure relevant microbiota nucleic acid instead of depleting abundant human nucleic acid (genomic DNA, ribosomal or mitochondrial RNA). Consequently, this hybridized approach can provide clinically relevant information from more cost effective and lower throughput sequencing techniques, important for decentralized testing and laboratories with limited resources. And although researchers have demonstrated that targeted hybridization-capture-based mNGS workflows for lower respiratory pathogen-specific sequences has critical implication [24], preceding or concurrent consideration of upper respiratory pathogenic infectivity and the potential trafficking effects on lower respiratory disease is less studied with this approach.

This is an important distinction from other amplicon sequencing methods laboratorians commonly use to analyze target specific genomic regions of respiratory microbiota—NGS of the 16S rRNA gene and target-agnostic mNGS. However, NGS of the 16S rRNA gene is limited to bacterial identification and is often restricted to the genus level [68]. Furthermore, the 16S rRNA-based approach does not provide crucial AMR marker detection for focused medication therapy [69]. Alternatively, the target-agnostic mNGS approach is becoming an increasingly viable technique for obtaining microbial nucleic acid sequence information for the diagnoses and treatment of pulmonary (co)infections [70]. Yet, this method is based on the depletion of host DNA, and deep sequencing of remaining DNA requires a higher depth because of a high degree of residual host DNA—requiring high capital investment (∼$1 million) in technology, bioinformatic specialization, and potential for turnaround lags because sample pooling is often needed to reduce run costs. As such, target-agnostic mNGS is most suitable in research laboratories or for epidemiological surveillance (not unlike the recent COVID-19 pandemic). Thereupon, these amplicon sequencing methods are largely unsuitable for deployment for routine diagnostic use, especially in decentralized or resource-limited clinical laboratories [29].

The ability of mNGS to outperform common diagnostic procedures in detecting unidentified pathogens has been reported. For example, [71] analyzed pathogens of respiratory infection by mNGS in bronchoalveolar lavage fluid (BALF) from immunocompetent pediatric patients with respiratory failure. The study reported significant identification of bacterial or viral sequencing reads in 8/10 patients not identified by conventional methods. Miao et al. [72] demonstrated that mNGS yielded higher specificity and sensitivity than microbial culture when identifying *Mycobacterium tuberculosis*, viruses, anaerobes, and fungi. Several other studies have reported more than 80% sensitivity of mNGS in detecting pathogens compared to traditional methods [73]. Moreover, co-infection is very common in clinical settings with studies reporting up to 70% of patients co-infected with bacteria-viruses, bacterial-fungal, or virus-fungal—with mNGS successfully identifying all co-infections compared to PCR analysis [73]. Likewise, our current study identified co-infection in (26/29) samples using the hybridization capture-based approach. This favors the advantage of using hybridization capture to identify a wide range of microorganisms in a single analysis. Overall, the hybridization capture-based mNGS workflow appears to be an emerging and promising technology for detecting respiratory pathogens more effectively, and with more clinical relevance, than conventional culture or PCR analysis.

Nevertheless, differentiating pathogens from commensal and colonized microorganisms is very challenging in respiratory (co)infections. Lungs in healthy and diseased individuals host different bacterial strains, and asymptomatic, or potentially pathogenic organisms are ubiquitously present in the lungs. For example, 20-50% of healthy airways are colonized by opportunistic bacteria such as *S. pneumoniae* and *H. influenza*—both considered to have high trafficking and lower respiratory infectivity potential. Although phenotypic grouping can aid the characterization of pathogenicity, other clinical factors should be considered for accurate diagnosis based upon data generated from a hybridization capture-based mNGS workflow—especially immunocompromised and immunosuppressed patients who are more vulnerable to (co)infections caused by a wide range of common and uncommon pathogens.

Finally, even though target-agnostic sequencing can identify the entire microbiome without *a priori* knowledge, the targeted hybridization-capture-based mNGS workflow is likely to have better application in clinical settings. This is because of the precise detection of clinically relevant organisms and better adaptability on low-throughput sequencing instruments. For example, using the hybridization-capture-based mNGS workflow, we correctly identified NATtrol™ Respiratory Panel 2.1 controls (ZeptoMetrix Inc. USA) from only 0.5 million reads. Indicating this approach can essentially analyze ∼24 samples with a small benchtop MiniSeq™ System, making it more easily adaptable and cost effective for the clinical setting. Importantly, there are no current Federal Drug Administration-approved devices or kits for sequencing infectious disease, requiring, in the United States, the extensive validation of the test’s accuracy, sensitivity, and specificity according to Clinical Laboratory Improvement Amendments (CLIA) guidelines.

## 5. Conclusions

Much of respiratory medicine is reliant on timely and precise diagnostics for critical treatment. But respiratory infections remain a leading cause of global mortality and morbidity despite advances in diagnosis and treatment. In this study (a) we compared nasopharyngeal samples from patients suspected of acute upper respiratory infection between a commercially available PCR assay with a targeted hybridization-capture-based mNGS workflow and (b) demonstrated that the hybridized approach may provide tremendous advantage in deciphering the etiological agent of respiratory (co)infections and provide clinical relance for trafficking potential. Important because the trafficking potential from the upper to the lower respiratory tract and infection severity depend on pathogen virulence, concomitant infections, and underlying respiratory comorbidities [74].

This is significant to respiratory medicine because this technology can be used to supplement current syndromic-based tests and data can quickly and effectively be phenotypically characterized for clinical (co)infection and comorbid consideration. This has significance for laboratory medicine because it demonstrated this approach can rapidly be interpreted with a user-friendly and reliable platform for collective intention without overburdening laboratory investments in technology and people [75,76]. Furthermore, this approach could be advanced into a pan-microbial diagnostic testing that utilizes a single workflow for all specimen types. Although we have demonstrated the analytical advantage of a targeted hybridization-capture-based mNGS workflow over targeted PCR analysis, further investigations are required to establish the clinical relevancy of phenotypic classifications and their value to trafficking predispositions and utility in respiratory medicine.

## Data Availability

All data produced in the present work are contained in the manuscript

## Supplementary Materials

The following supporting information can be downloaded at: www.mdpi.com/xxx/s1, Table S1-2.

## Author Contributions

Conceptualization, RC and RS; methodology, SA and RS; validation, SA, CR, and RS; data curation, SA, CR, AS, and VT; writing—original draft preparation, SA and RS; writing—review and editing, RC; supervision, RS; project administration, RS; All authors have read and agreed to the published version of the manuscript.

## Institutional Review Board Statement

This research used de-identified samples and institutional IRB exempted the study.

## Informed Consent Statement

Patient consent was not applicable due to research conducted on de-identified samples.

## Data Availability Statement

Harvard Dataverse

